# Associations of Chemical Exposures with Psychological Distress and Depression Diagnosis among Waste Pickers in Brasilia, Brazil: A Cross-Sectional Study

**DOI:** 10.64898/2026.06.19.26356069

**Authors:** Kaleigh L. King Stone, Sofia Pelagalli Maia, Allie Melanson, Makayla Steelman, Vanessa R.N. Cruvinel, Carla P. Marques, Nicole Macena, Steven Thygerson, Evan L. Thacker

**Author notes:** **Corresponding Author:** Kaleigh King Stone. **Author Contributions: King Stone, Kaleigh L:** Conceptualization, Formal Analysis, Methodology, Visualization, Writing - original draft, Project management. **Pelagalli Maia, Sofia:** Conceptualization, Formal Analysis, Methodology, Writing - Review & Editing. **Melanson, Allie:** Conceptualization, Formal Analysis, Methodology, Writing - Review & Editing. **Steelman, Makayla:** Conceptualization, Formal Analysis, Methodology, Writing - Review & Editing. **Cruvinel, Vanessa R.N.:** Data Curation, Resources, Writing - Review & Editing. **Marques, Carla P.:** Data Curation, Resources, Writing - Review & Editing. **Macena, Nicole:** Data Curation, Resources, Writing - Review & Editing. Thygerson, Steven: Writing - Review & Editing. **Thacker, Evan L:** Supervision, Writing - Review & Editing, Validation, Visualization.

## Abstract

**Introduction:** Waste pickers face chemical exposures. We evaluated whether chemical exposure is associated with psychological distress and depression.

**Methods:** A 2017 cross-sectional survey included 1,141 waste pickers working in the Estrutural open dump in Brasilia, Brazil. Participants self-reported occupational exposure to 11 chemical categories, 17 psychological distress symptoms, and depression diagnoses. Associations of chemical exposure with mean psychological distress scores and depression prevalence were assessed, adjusted for age, sex, marital status, and income.

**Results:** Mean psychological distress score was higher among those exposed to any chemical (mean of 8.1 vs 6.1; adjusted mean difference [aMD]: 1.8 [0.9, 2.7]) and higher among those exposed to each of 11 chemical categories, for example, smoke (aMD: 1.2 [0.6, 1.7]), batteries (aMD: 1.5 [1.0, 1.9], and oils (aMD: 1.3 [0.9, 1.8]). Depression was more prevalent among those exposed to oils (16.6% vs 10.6%; adjusted prevalence difference [aPD]: 6.3% [95% CI: 2.3, 10.2]), cleaning products (aPD: 5.4% [1.2, 9.5]), medications (aPD: 4.7% [0.6, 8.8]), and aerosols (aPD: 5.3% [1.3, 9.3]) but, not smoke, batteries, greases, insecticides, solvents, paints, chemical containers, or any chemical.

**Conclusion:** These associations highlight the need to consider policy level protections for waste pickers to reduce chemical exposure and guard against psychological distress. Further research is necessary to explore which specific chemicals, within broad chemical categories, are associated with psychological distress and depression.

## Introduction

It is estimated that in 2025 over 2 billion tons of municipal solid waste were generated globally, of which over 30% remains unmanaged [1]. Unmanaged waste poses serious environmental and public health risks, including the spread of infectious diseases, vector borne illnesses, respiratory illnesses from air pollution, and waterborne diseases [1,2]. In Brazil, municipal solid waste is placed into open-air dumps, which were set to terminate operations by 2022, in favor of waste management systems regulated by the municipalities [3,4]. Unfortunately, more than 50% of municipalities were unable to comply with this plan, leaving between two and three thousand unmanaged open dumps in Brazil [3,4]. While some open-air dumping sites are no longer operating in an official capacity, they remain in operation as unregulated or unofficial dumping sites (Zolnikov et al., 2018).

Given that a large proportion of the population’s waste remains inadequately managed by governmental bodies, it is important to recognize the role of waste pickers and the occupational challenges they face as informal workers. It is estimated that there are 15 million waste pickers in the world, 4 million in South America alone and nearly 230,000 in Brazil [5,6]. These waste pickers are often unorganized and autonomous individuals or groups who collect, sort, and sell recyclable materials from open-air landfills to earn their living (Zolnikov et al., 2018). While waste picking is officially recognized as an occupation in Brazil, a substantial proportion of waste pickers continue to work under informal and precarious conditions [7].

Waste pickers’ contributions to the ecosystem, economy, and public health are valuable, but their occupation makes them particularly vulnerable to a variety of health conditions and occupational hazards including exposure to household, commercial, and industrial chemicals [8]. In a study of 400 waste pickers living in Lucknow City, India, 63% indicated that they came into contact with decomposed materials and 72% reported contact with chemical substances including paints, sanitary napkins, household cleaning products, and biomedical waste [9]. Similarly, in a qualitative study across 9 dumping sites in South Africa, waste pickers reported exposure to dust, polluted air, aerosolized chemicals, decomposing materials, polluted water, and medical waste [10]. Hair and nail samples collected from 431 waste pickers working at the Estrutural dump in Brasília, Brazil, showed significantly higher concentrations of multiple heavy metals compared with less-exposed reference groups [11].

Chemical exposures may be associated with various mental health issues including psychological distress and depression. Psychological distress is the term for a “subjective sense of discomfort, mental anguish, perceived lack of control, anxiety, or stress” [12]. In a study among residents living in the Hebei Spirit oil spill accident area, on the coast of Taean, South Korea from 2009 to 2016, exposure to oil spills was associated with post-traumatic stress disorder (PTSD), depression, state anxiety, and trait anxiety [13]. Similarly, in a study among farmers in Austrailia, low-dose chronic exposure to agricultural pesticides was associated with poor mental health and suicide [14]. Among Japanese automotive workers in Kyushu, Japan, environmental and occupational exposures to chemicals were related to symptoms of mental illness, depressive feelings, memory disturbances, and anxiety [15].

Associations of chemical exposure and mental health outcomes among waste pickers is understudied. The lack of research combined with minimal regulations surrounding workplace protection may increase the risk of poor mental health outcomes such as depression and psychological distress. To address the gap, we examined the association of self-reported chemical exposures with self-reported psychological distress symptoms and diagnosis of depression among waste pickers in Brasilia, Brazil.

## Methods

### Study design, participants, and setting

In 2017, prior to the closure of the Estrutural open dump located in Brasilia, Brazil, 1,200 adult waste pickers, encompassing the entirety of the population, were registered in the Urban Cleaning Service in preparation for transition from the open dump to a recycling co-operative. All 1,200 individuals were invited to participate in the cross-sectional survey and 1,141 individuals (95%) agreed to participate and signed an informed consent [8]. For the present analysis, we excluded 14 individuals who reported in the survey not being a waste picker, leaving 1,127 (98.8%) for analysis.

### Chemical exposure measures

Exposures of interest were 11 dichotomous chemical category exposures including smoke, batteries, oils, greases, insecticides, solvents, paints, cleaning products, medications, aerosols, and chemical containers. Participants reported exposures to the chemical categories by answering the survey question “In the past month, have you been in contact with:” (Original Portuguese: “No último mês você teve contato com:”) and then selecting any of the chemical categories from a list on the survey. In addition to the 11 chemical categories, a participant was considered exposed to “any chemical” if they marked at least one category and not exposed to “any chemical” if they marked none of the 11 categories.

### Mental health measures

The outcomes of interest were psychological distress score and diagnosis of depression. Psychological distress score was calculated using a reduced Self-Report Questionnaire (SRQ-20), using a subset of 17 of the 20 original questions [16,17]. A Portuguese translation of the reduced SRQ-20 survey was used in the study; both Portuguese and English versions can be found in Supplemental Table 1 [16,17]. Depression was self-reported by answering “yes” to the survey question “Has a doctor or mental health professional ever diagnosed you with depression?” (Original Portuguese: “Algum médico ou profissional de saúde mental [Como psiquiatra ou psicólogo] já lhe deu o diagnóstico de depressão?”)

**Table 1.** Descriptive Epidemiology of Sociodemographic Characteristics.

| Sociodemographic Characteristics | Characteristics*<br>(Mean [SE] or N [%]) | Exposed to any<br>Chemical† (%) | Psychological Distress<br>Score (Mean [SE]) | Diagnosed with<br>Depression† (%) |
| --- | --- | --- | --- | --- |
| **Total Sample** | — | 94.5% | 8.0 (0.0) | 14.3% |
| **Age in years (Mean [SE])** | 40.6 (0.0) | — | --- | --- |
| **Sex** |  |  |  |  |
| Male | 370 (32.9%) | 92.9% | 5.8 (0.2) | 5.2% |
| Female | 757 (67.1%) | 95.3% | 9.0 (0.1) | 18.8% |
| **Race** |  |  |  |  |
| White | 133 (11.8%) | 96.3% | 7.5 (0.4) | 13.6% |
| Black/Asian/Indigenous | 291 (25.8%) | 94.0% | 8.2 (0.2) | 12.9% |
| Mixed | 703 (62.4%) | 94.4% | 8.0 (0.1) | 15.0% |
| **Marital Status** |  |  |  |  |
| Single | 692 (61.4%) | 94.2% | 8.1 (0.1) | 22.6% |
| Married/Long-term Partnership | 341 (30.2%) | 96.0% | 7.7 (0.2) | 11.7% |
| Divorced/Separated | 94 (8.4%) | 91.6% | 8.3 (0.4) | 14.4% |
| **Education** |  |  |  |  |
| None Completed | 417 (37.0%) | 94.4% | 8.0 (0.2) | 14.3% |
| Completed Elementary School | 519 (46.0%) | 94.7% | 8.1 (0.2) | 13.9% |
| Completed Middle School or Above | 191 (17.0%) | 94.2% | 7.6 (0.3) | 15.4% |
| **Individual Monthly Income in reals (R\$; Mean [SE])** | 524.9 (1.4) | — | — | — |

\*Percentages in this column are column percentages, counts were averaged across imputations and rounded to the nearest integer.
†Percentages in these columns are row percentages.

### Other measures

Participants self-reported their age (years), sex (male; female), race (white; black/Asian/indigenous; mixed), marital status (single, married/long-term partnership, divorced/separated), attained education (none; elementary school; middle school or above), and individual monthly income (reals [R$]).

### Multiple imputation

To increase precision and reduce bias by retaining all eligible participants in the data analysis instead of excluding participants with missing values, we conducted multiple imputation by chained equations [18]. Among eligible participants, 49.7% had at least one missing value for a mental health variable, chemical exposure variable, or other variable. Income was missing for 18.5% of participants; each other variable was missing for less than 3% of participants. The imputation model included each dichotomous chemical exposure variable, all 17 SRQ items, the dichotomous depression diagnosis, and all other measures listed above. We created 51 imputed datasets based on guidance in the literature to create at least as many imputations as the percentage of participants having at least one missing value [18].

### Statistical analysis

We summarized participant sociodemographic characteristics using means and standard errors for continuous measures and counts and percentages for categorical measures. For each chemical exposure in relation to mean psychological distress score, we calculated the difference in mean psychological distress score among those exposed to the chemical minus those unexposed to the chemical, using Poisson regression with an identity link. For each chemical exposure in relation to prevalence of depression, we calculated the difference in percent depressed among those exposed to the chemical minus those unexposed to the chemical, using linear regression with robust standard errors to adjust for sociodemographic characteristics. We adjusted all comparisons for age, sex, marital status, and individual monthly income. We did not adjust for race or education in our models because they were not strongly associated with chemical exposure, psychological distress, or depression in our sample. To assess potential effect measure modification by sex, we calculated sex-specific results in female and male subgroups, then tested the hypothesis of sex difference in the association of the chemical with the mental health outcome using the P value for the chemical×sex interaction term in the adjusted regression model. P values < 0.05 were considered statistically significant.

## Results

**Table 1** describes sociodemographic characteristics of the participants and the relationships of those characteristics with exposure to any chemical, mean psychological distress score, and prevalence of depression. Females had a higher proportion exposed to chemicals than males (95.3% vs 92.9%), higher mean psychological distress score (9.0 vs. 5.8), and higher prevalence of depression (18.8% vs. 5.2%),. Participants who indicated their marital status as single had a higher prevalence of depression (22.7%) compared to those who were married or in a long-term partnership (11.8%) or divorced (14.4%).

**Figure 1** and **Supplemental Table 2** examine unadjusted and adjusted associations between chemical category exposures and mean psychological distress score. Mean psychological distress score was statistically significantly higher for every chemical category exposure versus unexposed (all p < 0.001). For example, exposure to smoke was associated with a 1.2 point higher mean psychological distress score compared with people not exposed to smoke (95% CI: 0.6, 1.7; p < 0.001), and findings for all other chemical categories were similar.

**Figure 1:**
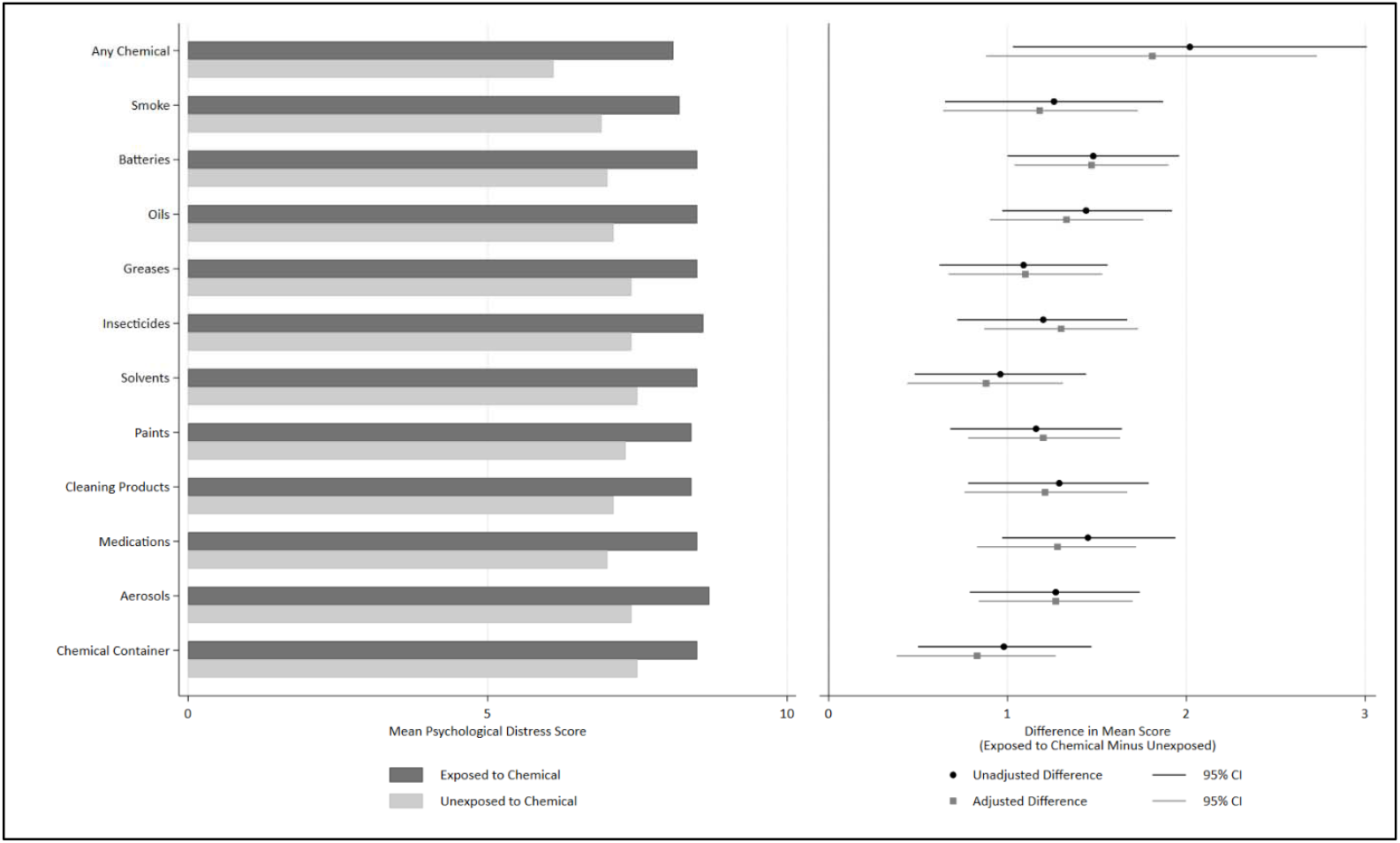
Associations between Chemical Exposures and Mean Psychological Distress Score. Caption: Figure 1 displays the mean psychological distress score among participants in our sample by each chemical exposure (left) and the difference in mean score in both the unadjusted and adjusted models (right).

**Figure 2** and **Supplemental Table 3** display unadjusted and adjusted associations between chemical category exposures and prevalence of depression. Prevalence of depression was higher for every chemical category exposure versus unexposed, with several statistically significant associations. For example, exposure to oils was significantly associated with having depression, with 6.3 more people diagnosed with depression per 100 people exposed to oils than among 100 unexposed to oils (95% CI: 2.3, 10.2; p = 0.002). Similarly, cleaning products (5.4 more cases per 100 [95% CI: 1.2, 9.5]; p = 0.012), medications (4.7 more cases per 100 [95% CI: 0.6, 8.8]; p = 0.012), and aerosols (5.3 more cases per 100 [95% CI: 1.3, 9.3]; p = 0.010) were significantly associated with a higher prevalence of depression. The other chemical categories were not significantly associated with depression.

**Figure 2:**
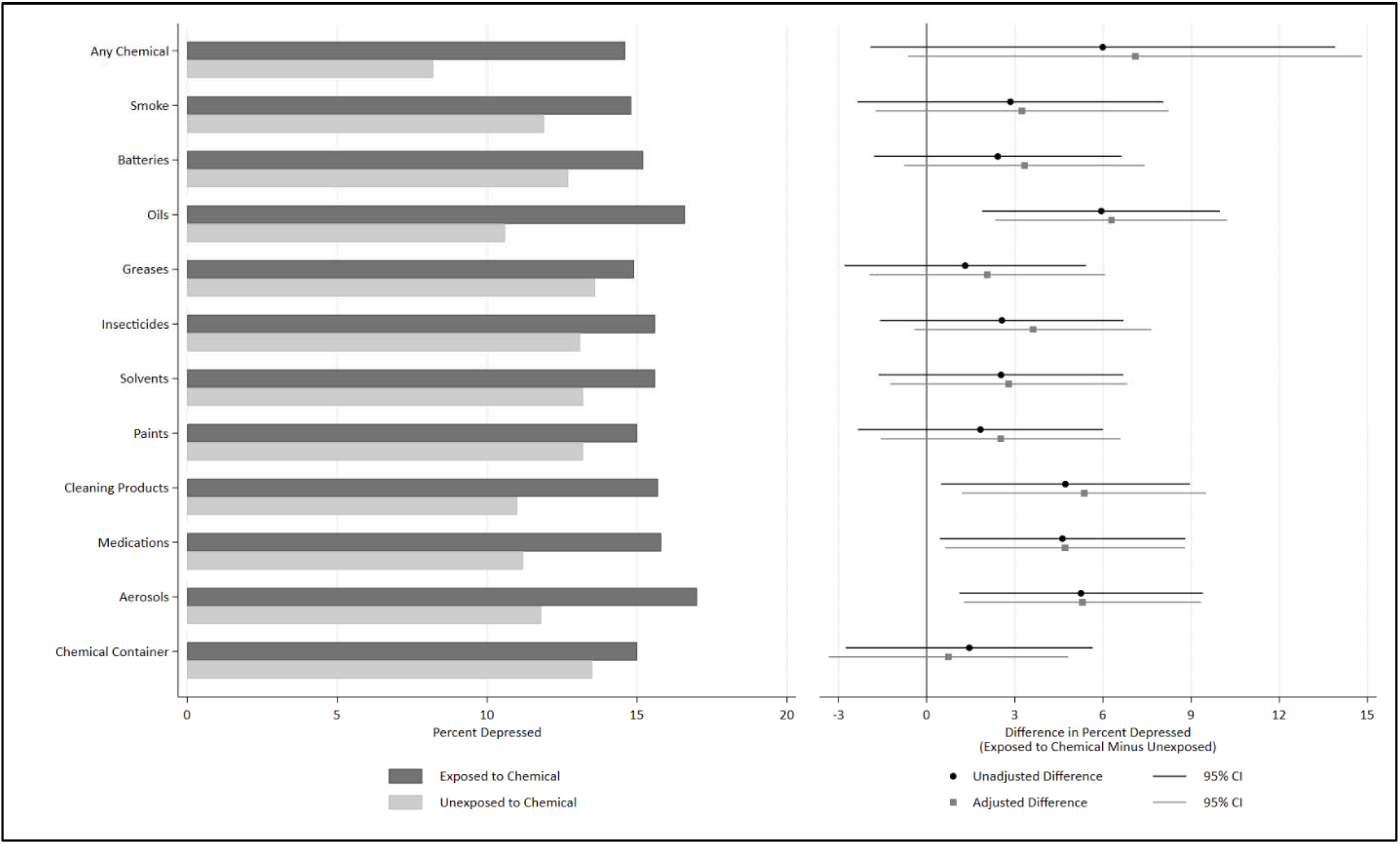
Associations between Chemical Exposures and Depression Prevalence per 100. Caption: Figure 2 displays the percent of participants in our sample who reported having ever been diagnosed with depression by a doctor (left) and the difference in the percent depressed when compared to the reference group in both the unadjusted and adjusted models (right).

**Table 4** illustrates effect measure modification by sex for associations of chemical category exposures with both the mean psychological distress score and depression prevalence. Estimated differences tended to be larger for females than for males, but most sex differences were not statistically significant, and precision of sex-specific estimates tended to be low. Exposure to aerosols was associated with depression among females (8.2 more cases per 100 females exposed to aerosols [95% CI: 2.6, 13.7]; p = 0.004) but not among males (0.5 more cases per 100 males exposed to aerosols [95% CI: −4.2, 5.2]; p = 0.826), which was a statistically significant sex difference (p for sex difference = 0.031).

**Table 4:**
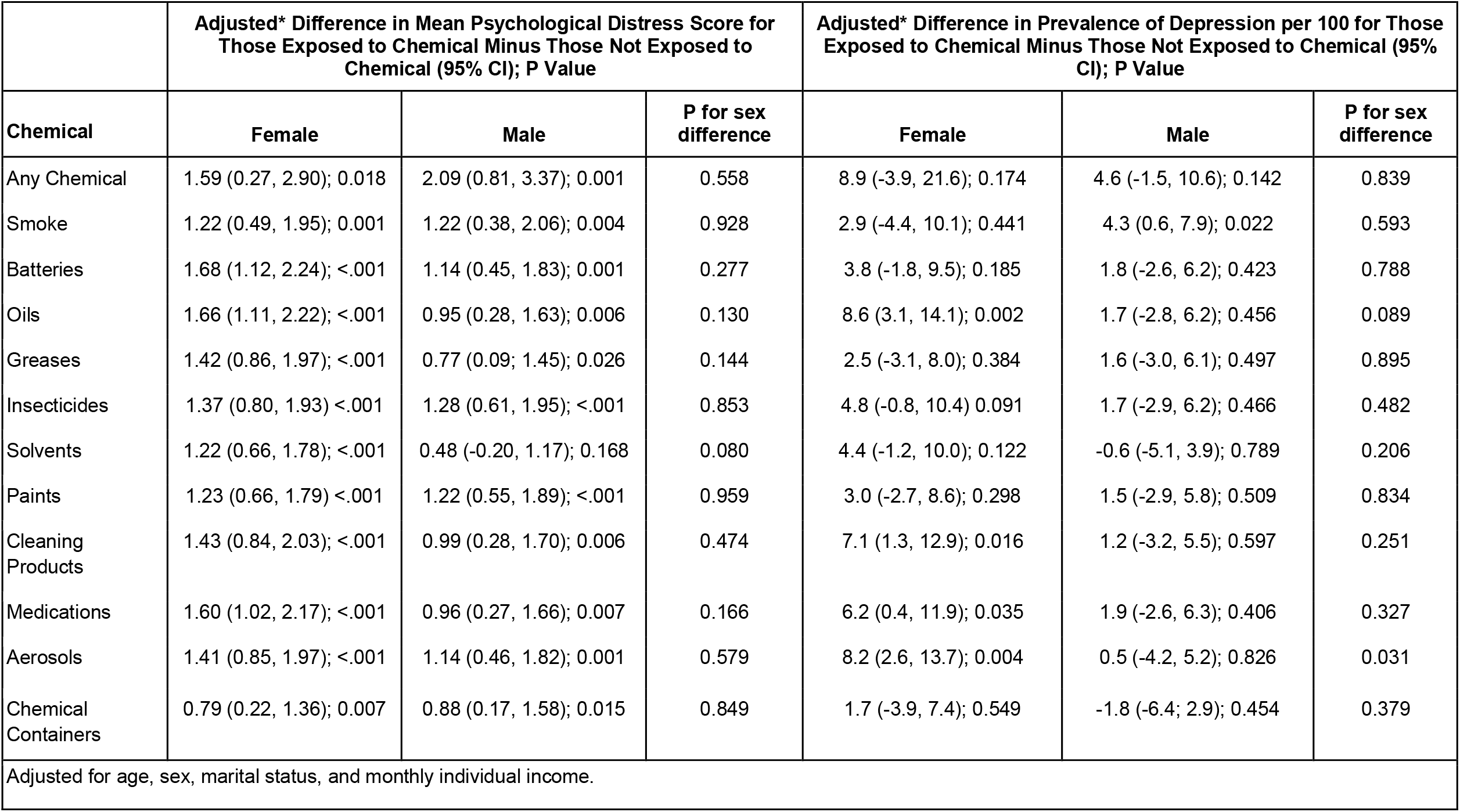
Effect Measure Modification by Sex for Associations between Chemical Exposure and Mean Psychological Distress Score and Depression Prevalence per 100.

|  | Adjusted* Difference in Mean Psychological Distress Score for Those Exposed to Chemical Minus Those Not Exposed to Chemical (95% CI); P Value |  |  | Adjusted* Difference in Prevalence of Depression per 100 for Those Exposed to Chemical Minus Those Not Exposed to Chemical (95% CI); P Value |  |  |
| --- | --- | --- | --- | --- | --- | --- |
| Chemical | Female | Male | P for sex difference | Female | Male | P for sex difference |
| Any Chemical | 1.59 (0.27, 2.90); 0.018 | 2.09 (0.81, 3.37); 0.001 | 0.558 | 8.9 (-3.9, 21.6); 0.174 | 4.6 (-1.5, 10.6); 0.142 | 0.839 |
| Smoke | 1.22 (0.49, 1.95); 0.001 | 1.22 (0.38, 2.06); 0.004 | 0.928 | 2.9 (-4.4, 10.1); 0.441 | 4.3 (0.6, 7.9); 0.022 | 0.593 |
| Batteries | 1.68 (1.12, 2.24); <.001 | 1.14 (0.45, 1.83); 0.001 | 0.277 | 3.8 (-1.8, 9.5); 0.185 | 1.8 (-2.6, 6.2); 0.423 | 0.788 |
| Oils | 1.66 (1.11, 2.22); <.001 | 0.95 (0.28, 1.63); 0.006 | 0.130 | 8.6 (3.1, 14.1); 0.002 | 1.7 (-2.8, 6.2); 0.456 | 0.089 |
| Greases | 1.42 (0.86, 1.97); <.001 | 0.77 (0.09, 1.45); 0.026 | 0.144 | 2.5 (-3.1, 8.0); 0.384 | 1.6 (-3.0, 6.1); 0.497 | 0.895 |
| Insecticides | 1.37 (0.80, 1.93) <.001 | 1.28 (0.61, 1.95); <.001 | 0.853 | 4.8 (-0.8, 10.4) 0.091 | 1.7 (-2.9, 6.2); 0.466 | 0.482 |
| Solvents | 1.22 (0.66, 1.78); <.001 | 0.48 (-0.20, 1.17); 0.168 | 0.080 | 4.4 (-1.2, 10.0); 0.122 | -0.6 (-5.1, 3.9); 0.789 | 0.206 |
| Paints | 1.23 (0.66, 1.79) <.001 | 1.22 (0.55, 1.89); <.001 | 0.959 | 3.0 (-2.7, 8.6); 0.298 | 1.5 (-2.9, 5.8); 0.509 | 0.834 |
| Cleaning Products | 1.43 (0.84, 2.03); <.001 | 0.99 (0.28, 1.70); 0.006 | 0.474 | 7.1 (1.3, 12.9); 0.016 | 1.2 (-3.2, 5.5); 0.597 | 0.251 |
| Medications | 1.60 (1.02, 2.17); <.001 | 0.96 (0.27, 1.66); 0.007 | 0.166 | 6.2 (0.4, 11.9); 0.035 | 1.9 (-2.6, 6.3); 0.406 | 0.327 |
| Aerosols | 1.41 (0.85, 1.97); <.001 | 1.14 (0.46, 1.82); 0.001 | 0.579 | 8.2 (2.6, 13.7); 0.004 | 0.5 (-4.2, 5.2); 0.826 | 0.031 |
| Chemical Containers | 0.79 (0.22, 1.36); 0.007 | 0.88 (0.17, 1.58); 0.015 | 0.849 | 1.7 (-3.9, 7.4); 0.549 | -1.8 (-6.4; 2.9); 0.454 | 0.379 |

Adjusted for age, sex, marital status, and monthly individual income.

## Discussion

This study demonstrated two main findings: first, association between all chemical category exposures and higher mean psychological distress symptoms, and second, associations between exposure to oils, cleaning products, medications, and aerosols with higher prevalence of diagnosed depression among waste pickers in Brasilia, Brazil. Although psychological distress and depression were substantially more common among females than among males, the estimated associations of chemical exposures with psychological distress and depression were of similar magnitude for males and females, except for aerosols having a stronger association with depression among females.

Because mental health challenges are more common among females than males, chemical exposure-related psychological distress and depression may pose a greater or more impactful burden on the female population than on the male population. Additional adverse circumstances may also contribute to the greater mental health burden in the female population. For example, in a qualitative study of 20 female waste pickers living in the Federal District of Brazil, females reported experiencing frequent abuse from partners and spouses as well as harassment from male waste pickers at work sites [19]. The impact of violence on females’ mental health may put them at increased risk for psychological distress and depression, increasing the burden of poor mental health among females when compared to males. Additional factors, including marital status, may also increase the burden of poor mental health among females.

Open dumps, such as Estrutural in Brasilia where the present study was conducted, have long been known to be dumping grounds for many chemical toxins. One study highlighted that waste pickers often face exposure to hazardous working conditions due to a lack of personal protective equipment and training on handling hazardous materials [20]. Another study of workers in three landfills in South Africa showed that waste pickers were commonly exposed to dust and other aerosolized pollutants across three landfills in South Africa [21]. There has been documented exposure of a variety of household chemicals, including electronic waste such as batteries, chlorines, pesticides and more among waste pickers [22]. The exposures noted in those previous studies are consistent with the types of chemical exposures reported at the Estrutural open dump in Brasilia, Brazil.

In a study of waste pickers, researchers suggested that the precarious working and living conditions may be a primary cause of psychological distress [23]. This hypothesis is supported by one study on first responders living in Washington state, in which researchers discovered that exposure to hazardous material was associated with higher psychological distress symptoms [24]. Similarly, household cleaning products like those commonly found in household waste, have also been linked to higher depression and anxiety. Additionally, aerosolized pollutants, specifically PM 2.5, are known to be associated with higher risk of depression [25]. In a study of 3427 adults from the National Health and Nutrition Examination Survey, researchers found that six out of the ten categories of environmental toxins tested, including polycyclic aromatic hydrocarbons (PAH) which occur as a byproduct of industrial processes including burning garbage, oils, and other organic substances, were associated with higher depressive symptoms [26].

Our study had a variety of strengths. First, nearly 100% of the population of waste pickers working in the Estrutural open dump were sampled for the survey, minimizing the possibility that selection bias affected the observed associations of chemical exposures with mental health. Second, psychological distress was measured with a version of the validated and widely used SRQ instrument. Third, multiple imputation was used to increase statistical power and reduce bias that could otherwise have arisen from excluding participants from analysis who had missing values for some variables. Fourth, we adjusted all comparisons for sociodemographic characteristics to reduce confounding. Fifth, we conducted subgroup analyses to explore and quantify sex differences in the observed associations.

Our study also had limitations. First, the participants all came from one dumping location, so the sample may not be representative of the population of waste pickers throughout Brazil or internationally. Second, chemical exposures and mental health outcomes were self-reported. Participants were asked to report exposure to chemicals within the last month, psychological distress symptoms within the last week, and depression diagnoses at any time in the past or at present, without using a validated diagnostic instrument for major depression, so incorrect recall may have led to misclassification, which may have attenuated observed associations. Third, due to the cross-sectional study design, we cannot be confident about the time sequence of chemical exposures preceding mental health outcomes versus mental health outcomes preceding chemical exposures. Fourth, because we studied broad chemical categories, we were unable to identify specific chemicals associated with mental health among waste pickers. Further studies would be needed to address specific chemicals in relation to psychological distress and depression.

### Implications of Findings

This study demonstrated associations of occupational chemical exposure with higher psychological distress and prevalence of depression among waste pickers. These associations highlight the need to consider policy-level occupational protections for workers in this vulnerable population to reduce chemical exposure, psychological distress, and negative mental health outcomes. Because our study examined broad categories of chemical exposure, not specific chemicals, future research is necessary to explore whether specific chemical exposures relate to psychological distress and depression. Additionally, further research is needed to establish causation and determine whether access to and use of personal protective equipment is effective in preventing chemical exposures and thereby reducing psychological distress and depression. Institutions and organizations working to support change in the lives of waste pickers should consider implementing regular mental health screenings and promoting access to mental health care.

### Declaration of generative AI and AI-assisted technologies in the manuscript preparation process

During the preparation of this work the authors used Chat GPT in order to resolve errors in SAS code and to improve the analytical process. After using this tool/service, the authors reviewed and edited the content as needed and take full responsibility for the content of the published article.

## Data Availability

The data analyzed in this study were provided by the original investigators and are not publicly available. De-identified data may be available from the corresponding author or the original investigators upon reasonable request and with appropriate permission.

## Acknowledgments

The authors wish to express their thanks to certain Brazilian institutions, without whose support it would not have been possible to carry out this study: the University of Brasília, where the Brazilian lecturers and researchers work; the partnership with School of Health Sciences (ESCS); the Government of the Federal District (GDF) and State Secretariat of Health of the Federal District, Brasilia (SESDF); the Primary Healthcare Center where the study was carried out, its managers and the professionals who participated; The collaborators of the “Stop, Think and Discard” project; The Federal District’s Urban Cleaning Service, Solid Waste Recycling Cooperatives, and the waste pickers who participated in the study.

## Statement of Ethics

The parent study received ethics approval from the Research Ethics Committee of Fundação de Ensino e Pesquisa em Ciências da Saúde (Opinion No. 1.598.531; CAAE 55754216.5.0000.5553). Participants signed an informed consent before data collection began. The present study involved secondary analysis of a fully de-identified dataset. According to institutional policy, analysis of de-identified data did not require IRB review.

## References

1 Kaza S, Yao L, Bhada-Tata P, et al. What a Waste 2.0. 2018. https://openknowledge.worldbank.org/entities/publication/d3f9d45e-115f-559b-b14f-28552410e90a (accessed 26 March 2026)

2 de Souza WM, Weaver SC. Effects of climate change and human activities on vector-borne diseases. Nat Rev Microbiol. 2024;22:476–91. doi: 10.1038/s41579-024-01026-0

3 Lino FAM, Ismail KAR, Castañeda-Ayarza JA. Municipal solid waste treatment in Brazil: A comprehensive review. Energy Nexus. 2023;11:100232. doi: 10.1016/j.nexus.2023.100232

4 Souza-Dal Bó GC de, Bernardin AM. The (de)valuation of solid waste in developing countries: The Brazilian case (2012–2022) after the national solid waste policy. Clean Waste Syst. 2026;13:100457. doi: 10.1016/j.clwas.2025.100457

5 Zolnikov TR, da Silva RC, Tuesta AA, et al. Ineffective waste site closures in Brazil: A systematic review on continuing health conditions and occupational hazards of waste collectors. Waste Manag. 2018;80:26–39. doi: 10.1016/j.wasman.2018.08.047

6 Bouvier M, Dias S. Statistical Brief No 29 Waste Pickers in Brazil: A Statistical Profile. 2021. https://www.wiego.org/wp-content/uploads/2021/12/WIEGO_Statistical_Brief_N29_Brazil_WPs.pdf (accessed 14 May 2026)

7 Valadão MAP, Silva RAD. National Solid Waste Policy: Analyzing the Collective and Diffuse Rights of Recyclable Materials Collectors. Ambiente Soc. 2024;27:e00111. doi: 10.1590/1809-4422asoc0111vu27l2oa

8 Martins GKM, Moraes Pintel Ramos H, Bashash M, et al. Assessment of occupational hazards facing waste pickers to support a proper closure of a large open dump. Waste Manag Res J Int Solid Wastes Public Clean Assoc ISWA. 2025;734242X251385835. doi: 10.1177/0734242X251385835

9 Kumari S, Kiran UV. Prevalence of health problems of rag pickers due to various hazards at Lucknow city. Hum Factors Healthc. 2022;2:100023. doi: 10.1016/j.hfh.2022.100023

10 Schenck CJ, Blaauw PF, Viljoen JM, et al. Exploring the Potential Health Risks Faced by Waste Pickers on Landfills in South Africa: A Socio-Ecological Perspective. Int J Environ Res Public Health. 2019;16:2059. doi: 10.3390/ijerph16112059

11 Rodrigues Gonçalves M, Cruvinel VRN, Verpaele S, et al. Metal levels in waste pickers in Brasilia, Brazil: hair and nail as exposure matrices: Journal of Toxicology and Environmental Health, Part A: Vol 87, No 2 - Get Access. 2023. https://www.tandfonline.com/doi/full/10.1080/15287394.2023.2276372 (accessed 26 March 2026)

12 CDC. About Mental Health. Ment. Health. 2025. https://www.cdc.gov/mental-health/about/index.html (accessed 22 April 2026)

13 Kim L, Huh D-A, Kang M-S, et al. Chemical exposure from the Hebei spirit oil spill accident and its long-term effects on mental health. Ecotoxicol Environ Saf. 2024;284:116938. doi: 10.1016/j.ecoenv.2024.116938

14 Khan N, Kennedy A, Cotton J, et al. A Pest to Mental Health? Exploring the Link between Exposure to Agrichemicals in Farmers and Mental Health. Int J Environ Res Public Health. 2019;16:1327. doi: 10.3390/ijerph16081327

15 Cui X, Lu X, Hisada A, et al. The correlation between mental health and multiple chemical sensitivity: a survey study in Japanese workers. Environ Health Prev Med. 2015;20:123–9. doi: 10.1007/s12199-014-0434-2

16 Beusenberg M, Orley JH, Organization WH. A User’s guide to the self reporting questionnaire (SRQ. World Health Organization 1994.

17 Mari JJ, Williams P. A validity study of a psychiatric screening questionnaire (SRQ-20) in primary care in the city of Sao Paulo. Br J Psychiatry J Ment Sci. 1986;148:23–6. doi: 10.1192/bjp.148.1.23

18 White IR, Royston P, Wood AM. Multiple imputation using chained equations: Issues and guidance for practice. Stat Med. 2011;30:377–99. doi: 10.1002/sim.4067

19 Santos JE dos, Dutra ABM, Cruvinel VRN, et al. VIOL ÊNCIA DOMÉSTICA E SOFRIMENTO PSíQUICO: NARRATIVAS DE MULHERES CATADORAS DE RESíDUOS SÓLIDOS. Uningá Rev. 2021;36:eURJ4053–eURJ4053. doi: 10.46311/2178-2571.36.eURJ4053

20 Imam ST, Rafizul IM. Occupational risks, vulnerabilities, and safety challenges among informal waste workers at the open disposal site in Khulna city. Int J Hyg Environ Health. 2025;266:114543. doi: 10.1016/j.ijheh.2025.114543

21 Uhunamure SE, Edokpayi JN, Shale K. Occupational Health Risk of Waste Pickers: A Case Study of Northern Region of South Africa - Uhunamure - 2021 - Journal of Environmental and Public Health - Wiley Online Library. 2021. https://onlinelibrary.wiley.com/doi/10.1155/2021/5530064 (accessed 26 March 2026)

22 Gutberlet J, Uddin SMN. Household waste and health risks affecting waste pickers and the environment in low- and middle-income countries. Int J Occup Environ Health. 2017;23:299–310. doi: 10.1080/10773525.2018.1484996

23 da Silva MC, Fassa AG, Kriebel D. Minor psychiatric disorders among Brazilian ragpickers: a cross-sectional study. Environ Health. 2006;5:17. doi: 10.1186/1476-069X-5-17

24 Kovalchick DF, Burgess JL, Kyes KB, et al. Psychological effects of hazardous materials exposures. Psychosom Med. 2002;64:841–6. doi: 10.1097/01.psy.0000021947.90613.92

25 Deng Y, Hao H, Zhu Q, et al. Exposure to Multiple Fine Particulate Matter Components and Incident Depression in the US Medicare Population. JAMA Netw Open. 2025;8:e2551042. doi: 10.1001/jamanetworkopen.2025.51042

26 Guo J, Garshick E, Si F, et al. Environmental Toxicant Exposure and Depressive Symptoms. JAMA Netw Open. 2024;7:e2420259. doi: 10.1001/jamanetworkopen.2024.20259

