## Supplemental Table 1; Supplemental Table 2; Supplemental Table 3 for "Associations of Chemical Exposures with Psychological Distress and Depression Diagnosis among Waste Pickers in Brasilia, Brazil: A Cross-Sectional Study"

| **Supplemental Table 1: Self-Report Questionnaire (SRQ) English and Portuguese** | | |
| --- | --- | --- |
|  | **English** | **Portuguese** |
| **SRQ-1** | Do you often have headaches? | Você tem dores de cabeça freqüente? |
| **SRQ-2** | Is your appetite poor? | Tem falta de apetite? |
| **SRQ-3** | Do you sleep badly? | Dorme al? |
| **SRQ-4** | Are you easily frightened? | Assusta-se com facilidade? |
| **SRQ-5** | Do your hands shake? | Tem tremores nas mãos? |
| **SRQ-6** | Do you feel nervous, tense or worried? | Sente-se nervoso(a), tenso(a) ou preocupado(a) |
| **SRQ-7** | Is your digestion poor? | Tem má digestão? |
| **SRQ-8** | Do you feel unhappy? | Tem se sentido triste ultimamente? |
| **SRQ-9** | Do you cry more than usual? | Tem chorado mais do que de costume? |
| **SRQ-10** | Do you find it difficult to enjoy your daily activities? | Encontra dificuldades para realizar com satisfação suas atividades diárias? |
| **SRQ-11** | Do you find it difficult to make decisions? | Tem dificuldades para tomar decisões? |
| **SRQ-12** | Is your daily work suffering? | Tem dificuldades no serviço (seu trabalho é penoso, causa-lhe sofrimento?) |
| **SRQ-13** | Are you unable to play a useful part in life? | É incapaz de desempenhar um papel útil em sua vida? |
| **SRQ-14** | Have you lost interest in things? | Tem perdido o interesse pelas coisas? |
| **SRQ-15** | Do you feel that you are a worthless person? | Você se sente uma pessoa inútil, sem préstimo? |
| **SRQ-16** | Has the thought of ending your life been on your mind? | Tem tido idéia de acabar com a vida? |
| **SRQ-17** | Do you feel tired all the time? | Sente-se cansado(a) o tempo todo? |

| **Supplemental Table 2: Associations between Chemical Exposures and Mean Psychological Distress Score** | | | | |
| --- | --- | --- | --- | --- |
|  | **Psychological Distress Score (Mean [SE])** | | **Difference in Mean Psychological Distress Score for Those Exposed to Chemical Minus Not Exposed to Chemical (95% CI); P Value** | |
| **Chemical** | **Exposed to Chemical** | **Not Exposed to Chemical** | **Unadjusted** | **Adjusted*** |
| Any Chemical | 8.1 (0.1) | 6.1 (0.6) | 2.0 (1.0, 3.0); <0.001 | 1.8 (0.9, 2.7); <0.001 |
| Smoke | 8.2 (0.1) | 6.9 (0.3) | 1.3 (0.7, 1.9); <0.001 | 1.2 (0.6, 1.7); <0.001 |
| Batteries | 8.5 (0.1) | 7.0 (0.2) | 1.5 (1.0, 2.0); <0.001 | 1.5 (1.0, 1.9); <0.001 |
| Oils | 8.5 (0.1) | 7.1 (0.2) | 1.4 (1.0, 1.9); <0.001 | 1.3 (0.9, 1.8); <0.001 |
| Greases | 8.5 (0.2) | 7.4 (0.2) | 1.1 (0.6, 1.6); <0.001 | 1.1 (0.7, 1.5); <0.001 |
| Insecticides | 8.6 (0.2) | 7.4 (0.2) | 1.2 (0.7, 1.7); <0.001 | 1.3 (0.9, 1.7); <0.001 |
| Solvents | 8.5 (0.2) | 7.5 (0.2) | 1.0 (0.5, 1.4); <0.001 | 0.9 (0.4, 1.3); <0.001 |
| Paints | 8.4 (0.1) | 7.3 (0.2) | 1.2 (0.7; 1.6); <0.001 | 1.2 (0.8, 1.6); <0.001 |
| Cleaning Products | 8.4 (0.1) | 7.1 (0.2) | 1.3 (0.8, 1.8); <0.001 | 1.2 (0.8, 1.7); <0.001 |
| Medications | 8.5 (0.1) | 7.0 (0.2) | 1.5 (1.0, 2.0); <0.001 | 1.3 (0.8, 1.7); <0.001 |
| Aerosols | 8.7 (0.2) | 7.4 (0.2) | 1.3 (0.8; 1.7); <0.001 | 1.3 (0.8, 1.7); <0.001 |
| Chemical Containers | 8.5 (0.2) | 7.5 (0.2) | 1.0 (0.5, 1.5); <0.001 | 0.8 (0.4, 1.3); <0.001 |
| *Adjusted for age, sex, marital status, and monthly individual income. | | | | |

| **Supplemental Table 3. Associations between Chemical Exposures and Depression Prevalence per 100** | | | | | | | | |
| --- | --- | --- | --- | --- | --- | --- | --- | --- |
|  | **Exposed to chemical* (N)** | | **Not Exposed to Chemical* (N)** | | **Prevalence of Depression† (%)** | | **Difference in Prevalence of Depression per 100 for Those Exposed to Chemical Minus Not Exposed to Chemical (95% CI); P Value** | |
| **Chemical** | **Depressed** | **Not depressed** | **Depressed** | **Not depressed** | **Exposed to Chemical** | **Not Exposed to Chemical** | **Unadjusted** | **Adjusted‡** |
| Any Chemical | 156 | 909 | 5 | 56 | 14.6% | 8.2% | 6.0 (-1.9, 13.9); 0.138 | 7.1 (-0.6, 14.8); 0.072 |
| Smoke | 139 | 803 | 22 | 163 | 14.8% | 11.9% | 2.9 (-2.4, 8.1); 0.284 | 3.2 (-1.7, 8.2); 0.202 |
| Batteries | 110 | 616 | 51 | 350 | 15.2% | 12.7% | 2.4 (-1.8, 6.6); 0.260 | 3.3 (-0.8, 7.4); 0.111 |
| Oils | 115 | 579 | 46 | 387 | 16.6% | 10.6% | 5.9 (1.9, 10.0); 0.004 | 6.3 (2.3, 10.2); 0.002 |
| Greases | 88 | 503 | 73 | 463 | 14.9% | 13.6% | 1.3 (-2.8, 5.4); 0.532 | 2.1 (-1.9, 6.1); 0.313 |
| Insecticides | 82 | 442 | 79 | 524 | 15.6% | 13.1% | 2.6 (-1.6, 6.7); 0.227 | 3.6 (-0.4, 7.7); 0.079 |
| Solvents | 80 | 432 | 81 | 534 | 15.6% | 13.2% | 2.5 (-1.6, 6.7); 0.234 | 2.8 (-1.3, 6.8); 0.176 |
| Paints | 103 | 584 | 58 | 382 | 15.0% | 13.2% | 1.8 (-2.3,6.0); 0.389 | 2.5 (-1.6, 6.6); 0.226 |
| Cleaning Products | 124 | 667 | 37 | 299 | 15.7% | 11.0% | 4.7 (0.5, 9.0); 0.029 | 5.4 (1.2, 9.5); 0.012 |
| Medications | 120 | 641 | 41 | 325 | 15.8% | 11.2% | 4.6 (0.5, 8.8); 0.030 | 4.7 (0.6, 8.8); 0.024 |
| Aerosols | 91 | 443 | 70 | 523 | 17.0% | 11.8% | 5.3 (1.1, 9.4); 0.013 | 5.3 (1.3, 9.3); 0.010 |
| Chemical Containers | 85 | 480 | 76 | 486 | 15.0% | 13.5% | 1.5 (-2.8, 5.7); 0.500 | 0.7 (-3.3, 4.8);0.722 |
| *Counts were averaged across imputations and rounded to the nearest integer.  †Percentages are row percentages.  ‡Adjusted for age, sex, marital status, and monthly individual income. | | | | | | | | |
